# Quantitative analysis of SARS-CoV-2 RNA from wastewater solids in communities with low COVID-19 incidence and prevalence

**DOI:** 10.1101/2020.08.11.20173062

**Authors:** Patrick M. D’Aoust, Élisabeth Mercier, Danika Montpetit, Jian-Jun Jia, Ilya Alexandrov, Nafisa Neault, Aiman Tariq Baig, Janice Mayne, Xu Zhang, Tommy Alain, Mark R. Servos, Malcolm MacKenzie, Daniel Figeys, Alex E. MacKenzie, Tyson E. Graber, Robert Delatolla

## Abstract

In the absence of an effective vaccine to prevent COVID-19 it is important to be able to track community infections to inform public health interventions aimed at reducing the spread and therefore reduce pressures on health-care units, improve health outcomes and reduce economic uncertainty. Wastewater surveillance has rapidly emerged as a potential tool to effectively monitor community infections for severe acute respiratory syndrome coronavirus 2 (SARS-CoV-2), through measuring trends of viral RNA signal in wastewater systems. In this study SARS-CoV-2 viral RNA N1 and N2 genes are quantified in solids collected from influent post grit solids (PGS) and primary clarified sludge (PCS) in two water resource recovery facilities (WRRF) serving Canada’s national capital region, i.e., the City of Ottawa, ON (pop. ≈ 1.1M) and the City of Gatineau, QC (pop. ≈ 280K). PCS samples show signal inhibition using RT-ddPCR compared to RT-qPCR, with PGS samples showing similar quantifiable concentrations of RNA using both assays. RT-qPCR shows higher frequency of detection of N1 and N2 genes in PCS (92.7, 90.6%) as compared to PGS samples (79.2, 82.3%). Sampling of PCS may therefore be an effective approach for SARS-CoV-2 viral quantification, especially during periods of declining and low COVID-19 incidence in the community. The pepper mild mottle virus (PMMV) is determined to have a less variable RNA signal in PCS over a three month period for two WRRFs, regardless of environmental conditions, compared to *Bacteroides* 16S rRNA or human eukaryotic 18S rRNA, making PMMV a potentially useful biomarker for normalization of SARS-CoV-2 signal. PMMV-normalized PCS RNA signal from WRRFs of two cities correlated with the regional public health epidemiological metrics, identifying PCS normalized to a fecal indicator (PMMV) as a potentially effective tool for monitoring trends during decreasing and low-incidence of infection of SARS-Cov-2 in communities.

## 1. Introduction

Since the onset of the novel coronavirus disease in 2019 (COVID-19), the rapid transmission and global spread of the disease has placed significant strain on public health agencies around the world. Detection of SARS-CoV-2 RNA in nasopharyngeal (NP) swab specimens by reverse transcription quantitative polymerase chain reaction (RT-qPCR) is the standard diagnostic test to confirm COVID-19. Accurately measuring the prevalence of COVID-19 in many countries has been complicated by limited and/or biased NP testing (targeting symptomatic groups) and an asymptomatic, or mildly symptomatic infectious period in a significant proportion of cases (Long et al., 2020; Pan et al., 2020). Additional detection tools are thus desirable to mitigate these challenges and provide public health agencies and governments new metrics to help guide their implementation of societal restrictions (Daughton, 2009; Hill et al., 2020; Thompson et al., 2020).

Recent systematic reviews and meta-analyses of the current peer-reviewed and preprint literature confirm fecal SARS-CoV-2 viral RNA detection in roughly half of COVID-19 patients (Gupta et al., 2020; Parasa et al., 2020). Moreover, a systematic review and meta-analysis of SARS-CoV-2 viral RNA detection profiles in several different types of COVID-19 patient specimens found that positive detection rates were higher in rectal and sputum swabs than in the commonly used NP swab (Bwire et al., 2020). These data provide a clear rationale to probe wastewater for SARS-CoV-2 RNA.

Medema et al. (2020) first reported the detection of SARS-CoV-2 viral RNA in wastewater from WRRFs located in seven different cities in the Netherlands. SARS-CoV-2 viral RNA has subsequently been identified and is being monitored at numerous WRRFs around the world (Ahmed et al., 2020a; Alpaslan-Kocamemi et al., 2020; Bar Or et al., 2020; Haramoto et al., 2020; La Rosa et al., 2020; Medema et al., 2020; Nemudryi et al., 2020; Peccia et al., 2020a; Randazzo et al., 2020; Rimoldi et al., 2020; Wu et al., 2020; Wurtzer et al., 2020; Zhang et al., 2020) The successful monitoring of the viral signal has led the Netherlands (National Institute for Public Health and the Environment, 2020), Australia (Dalzell, 2020), Germany (Pleitgen, 2020) and Finland (Yle, 2020) to plan and implement national wastewater surveillance programs for SARS-CoV-2 as a viral tracking tool to complement existing public health metrics. There are also early and promising indications from several research groups that wastewater surveillance of SARS-CoV-2 might be predictive, providing earlier warning of community outbreak than current NP-based PCR diagnostics.

Although studies reported some success in the detection and even quantitation of SARS-CoV-2 RNA by RT-qPCR in wastewaters over the course of community COVID-19 outbreaks, poor assay sensitivity and systematic variation represent significant challenges, particularly in regions with low COVID-19 prevalence (Bar-On et al., 2020; Michael-Kordatou et al., 2020; Orive et al., 2020; Randazzo et al., 2020). Specifically, monitoring in communities with low incidence has demonstrated high PCR Ct values and hence variable or unquantifiable data being collected due to very low concentrations of the viral fragments in wastewaters. In this regard, at least two groups have identified improved sensitivity in solids-rich wastewater samples collected from WRRFs in communities with low incidence and prevalence (<25 active cases/100,000 population) (Balboa et al., 2020; Peccia et al., 2020b, 2020a). However, it has been observed that due to variations both in case numbers and influent wastewater sample data (Medema et al., 2020; Wu et al., 2020), studies have so far reported high day to day variance and noise (Balboa et al., 2020; Peccia et al., 2020a); which is a key challenge in establishing trends and extracting meaningful information from SARS-CoV-2 wastewater sentinel surveillance programs to date. This study investigates and optimizes the detection of SARS-CoV-2 RNA in wastewater influent solids (post-grit solids; PGS) and primary clarified sludge (PCS) in two municipal WRRFs serving Ottawa and Gatineau beginning after the height of the epidemic with a period (April to May 2020) characterized by decreasing COVID-19 incidence and a subsequent period (May to June 2020) of low COVID-19 prevalence. Using both RT-qPCR and RT-droplet digital (dd) PCR, rigorous quality control metrics are applied to compare the detection sensitivity of viral N1 and N2 RNA in PGS compared to PCS using two different established primer/probe sets. Furthermore, the study tests the human microbiome-specific HF183 *Bacteroides* 16S ribosomal RNA (rRNA) the eukaryotic 18S rRNA and pepper mild mottle virus (PMMV) RNA as reliable and robust nucleic acid normalization biomarkers that can be used to control systematic noise associated with variances in WRRF daily operations, sampling, storage, processing and analysis of the samples. Finally, the study compares and correlates biomarker normalized longitudinal data sets of the two municipalities with epidemiological metrics to evaluate the usefulness of SARS-CoV-2 viral measurements in wastewater as a complimentary tool to clinical testing in a community during decreasing and low COVID-19 incidence.

## Materials and methods

### 2.1. Characteristics of the City of Ottawa and Gatineau WRRFs

Post-grit chamber influent solids and primary clarified sludge samples were collected from the City of Ottawa’s Robert O. Pickard Environmental Centre, Ontario and the City of Gatineau, Quebec water resource recovery facilities (WRRFs). The two facilities are located across the Ottawa River from each other in the national capital region of Canada (Figure 1). The two WRRFs service over 1.3 million people, or approximately 3.7% of Canada’s total population. The sewershed of the City of Ottawa WRRF services approximately 1.1M people and the sewershed of the city of Gatineau WRRF services approximately 280K people.

**Figure 1.**
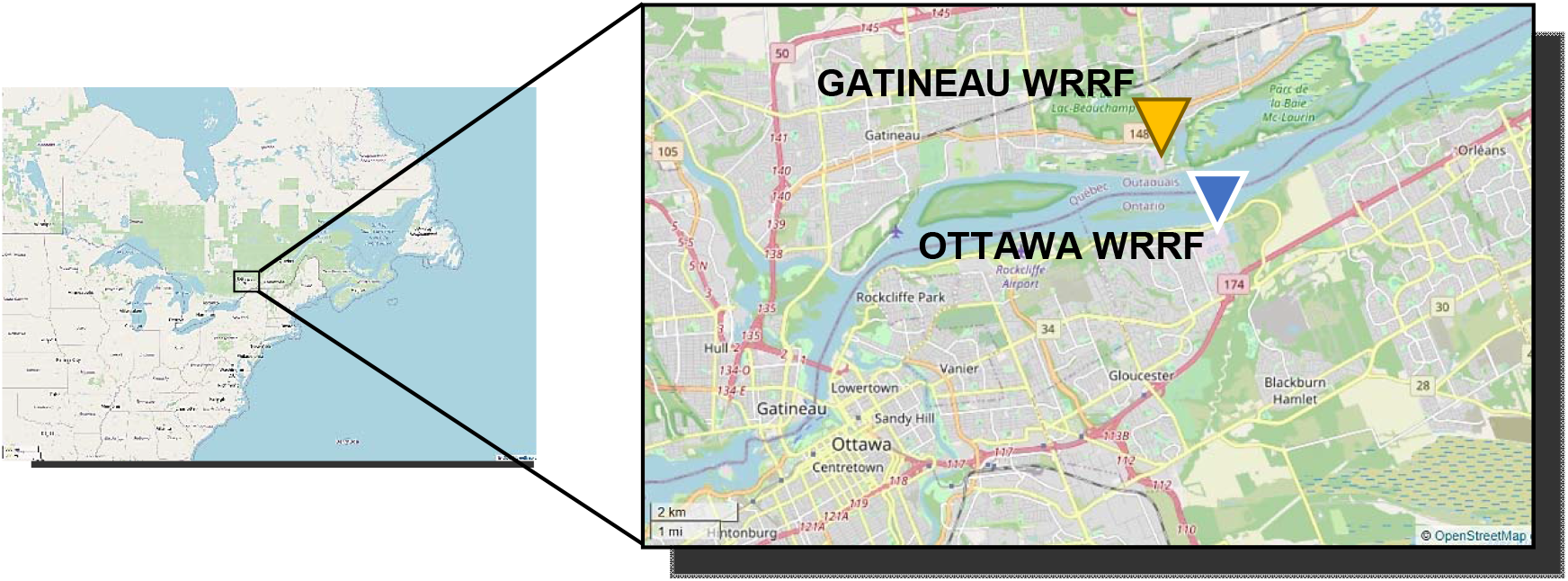
Location of the city of Ottawa and Gatineau WRRFs.

Ottawa and Gatineau WRRFs are designed and operated as conventional activated sludge treatment systems (Table 1). The grit chambers of both facilities, where a portion of the samples are collected in this study, are located toward the front of both WRRF treatment trains and are fed by coarse and fine screened wastewaters. The grit chambers of both facilities subsequently feed the primary clarifiers, where remaining portion of the samples are collected in this study. The hydraulic residence time of the Ottawa sewershed ranges from 2 hours to 35 hours, with an average residence time of approximately 12 hours. In comparison, the hydraulic residence time of Gatineau’s sewershed ranges from 2 hours to 7 hours, with an average of approximately 4 hours.

**Table 1:**
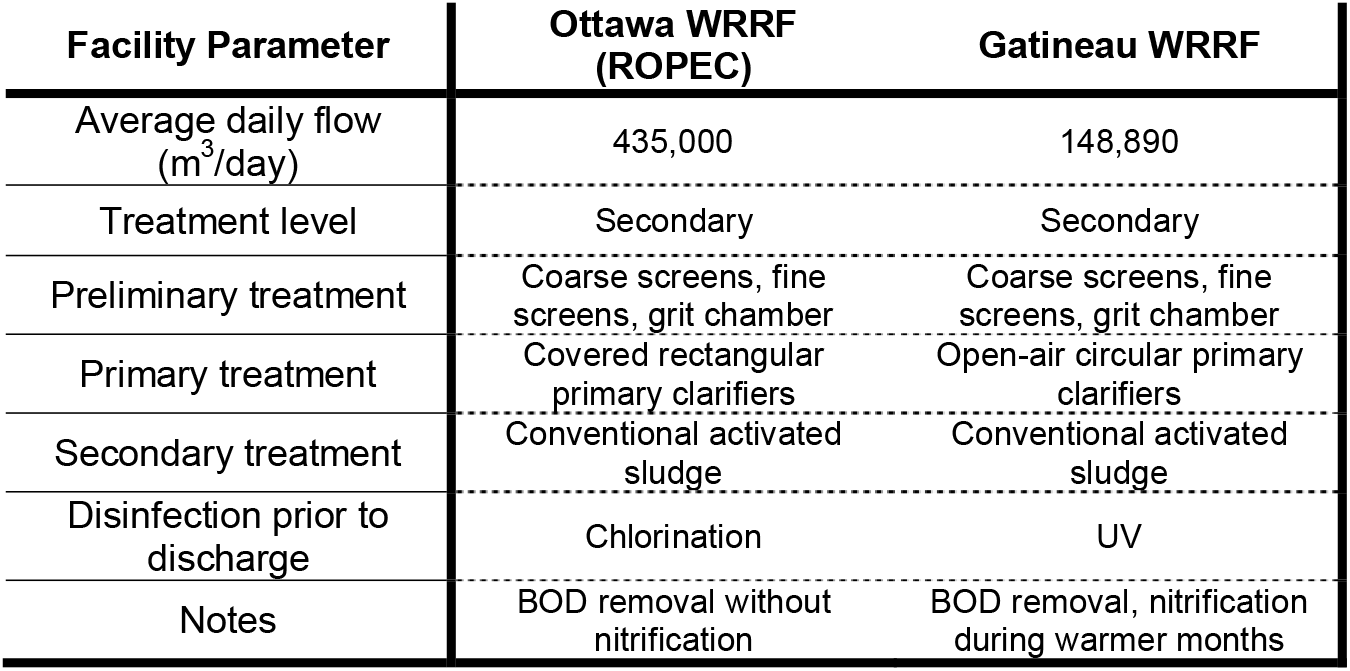
Characteristics of surveyed WRRFs

| Facility Parameter | Ottawa WRRF (ROPEC) | Gatineau WRRF |
| --- | --- | --- |
| Average daily flow (m<sup>3</sup>/day) | 435,000 | 148,890 |
| Treatment level | Secondary | Secondary |
| Preliminary treatment | Coarse screens, fine screens, grit chamber | Coarse screens, fine screens, grit chamber |
| Primary treatment | Covered rectangular primary clarifiers | Open-air circular primary clarifiers |
| Secondary treatment | Conventional activated sludge | Conventional activated sludge |
| Disinfection prior to discharge | Chlorination | UV |
| Notes | BOD removal without nitrification | BOD removal, nitrification during warmer months |

### 2.2. Wastewater sampling and analysis

#### 2.2.1. PGS samples

Fourteen and nine 24-hour composite PGS samples were analyzed from the Ottawa and Gatineau WRRFs, respectively. Clean 250 mL HDPE sampling bottles were sanitized with a 10% bleach solution and then washed with RNAse AWAY™ (ThermoFisher, Ottawa, Canada), rinsed with deionized water, and sealed. The PGS samples in this study are collected in the stream exiting the grit chambers. These samples have large debris removed via screens as was the dense grit via the grit chamber. A bio-banked wastewater influent sample from a nearby WRRF collected in August 2019 was utilized as a SARS-COV-2 negative control (Supplemental Figure S1).

250 mL hourly composite samples were collected over a 24-hour period (for a total of 6 L) using an ISCO autosampler (Hoskin Scientific, Burlington, Canada) that collects directly from the exit stream of the grit chamber units at both facilities. The samples in the ISCO autosamplers were maintained at approximately 4°C during sampling with the frequent addition of ice (with a maximum recorded temperature of 7°C across the study). Starting in June, the ISCO autosamplers were linked to refrigerators, allowing the samples to be kept at temperatures of approximately 2°C immediately upon collection. The harvested samples were transported from the WRRFs to the laboratory in coolers packed on ice and were immediately refrigerated at 4°C for a maximum of 24 hours prior to analysis.

#### 2.2.2. PCS samples

For the first 55 days of the study, grab samples of PCS were collected every second week at the Ottawa and Gatineau WRRFs. These sludge samples were harvested from the primary sludge streams in the two facilities at the manifold where the primary clarified sludge that exits all primary clarifiers was mixed into a single stream. From day 56 onward, with the stronger RNA signal detected in PCS samples as compared to PGS samples, 24-hour composite PCS samples were collected by plant process technicians every other day at the Ottawa facility. The 24-hour composite samples collected at the Ottawa WRRF were comprised of four grab samples collected every 6 hours. Upon collection, samples were stored on-site at the Ottawa facility at 4°C in a refrigerator until mixed to form daily 24-hour composite samples and transported on ice to the laboratory the subsequent day. All samples were stored at 4°C at the laboratory and processed within 6 hours of arrival. Samples which could not immediately be analyzed were stored at 4°C for a maximum of 24 hours prior to analysis in the laboratory. Meanwhile, in Gatineau, an ISCO autosampler was linked to a refrigerator and was connected to a PCS sampling port. The autosampler collected hourly grab samples of 250 mL, which were subsequently mixed to form a 24-hour composite sample. Due to the size differences of the two facilities in this study and the available resources at the two facilities during the COVID-19 pandemic in Canada, the Gatineau facility sampling frequency was limited to a maximum of once a week as opposed to every second day as was performed at the larger Ottawa facility. The 24-hour composite samples were collected and transported on ice to the laboratory as outlined for the Ottawa samples.

#### 2.2.3. Wastewater quality characterization of samples

The following PGS and PCS sample wastewater quality constituents were analyzed upon collection: biological oxygen demand (BOD) (5210 B) (APHA, WEF, 2012), chemical oxygen demand (COD) (SM 5220 D) (APHA, WEF, 2012), total suspended solids, volatile suspended solids, total solids and total volatile solids (TSS, VSS, TS & VS) (SM 2540 D, E & B) (APHA, WEF, 2012), total ammonia nitrogen (TAN) (SM 4500-C) (APHA, 1989). Dissolved oxygen (DO) and pH values were measured onsite during collection of samples with a YSI ProODO (Yellow Springs, FL) and HACH PHC201/HACH HQ40d probe/meter combo (Loveland, CO).

### 2.3. SARS-CoV-2 concentration

A preliminary study was first performed on partitioned 24-hour composite PGS samples to identify fractions with SARS-CoV-2 RNA positivity (Figure 2). The 6 L, 24-hour composite PGS samples were first settled at 4°C for an hour. The supernatant was subsequently decanted and serially filtered through a 1.5 μm glass fiber filter (GFF) followed by a 0.45 μm GF6 mixed cellulose ester (MCE) filter (filtrate fraction). An eluate fraction was then collected by passing 32 mL of elution buffer (0.05 M KH_2_PO_4_, 1.0 M NaCl, 0.1 % (v/v) Triton X-100, pH 9.2) through the spent filters. Each of the three fractions were subsequently PEG-concentrated and extracted and analyzed for SARS-CoV-2 RNA.

**Figure 2:**
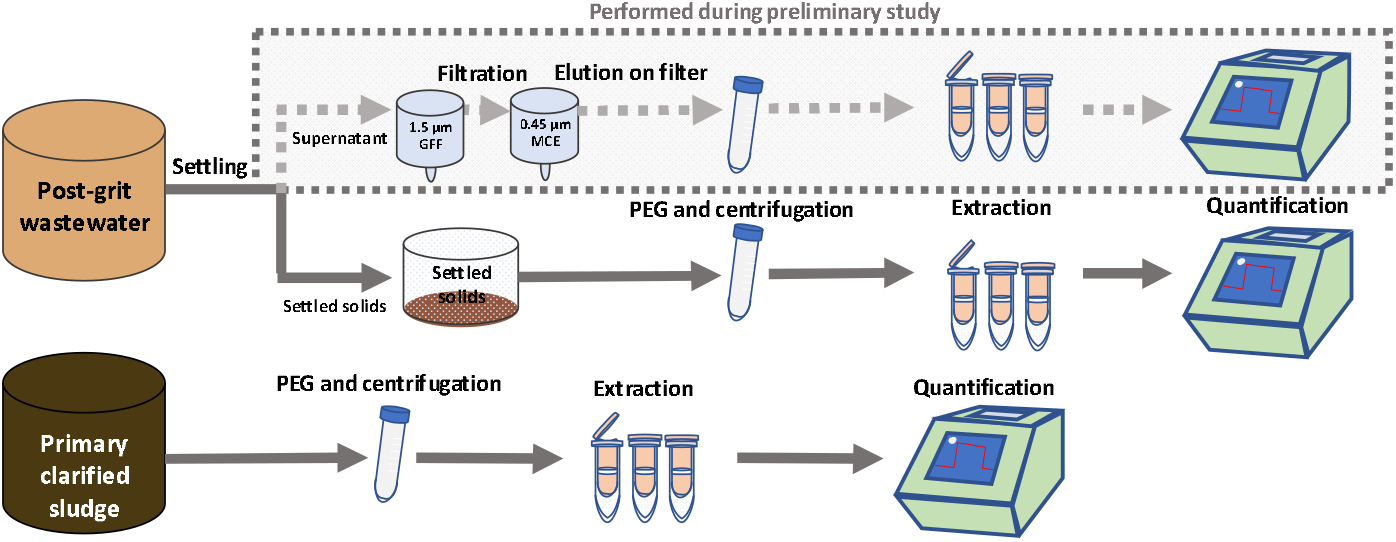
Flowchart showing sample work-up and processing for PGS and PCS, including RNA concentration, extraction and quantification.

To concentrate viral particles, nucleic acids, and proteins, 32 mL of PGS or PCS was precipitated with polyethylene glycol (PEG) 8000 at a final concentration of 80 g/L and 0.3M g/L NaCl, pH 7.3 and in a final volume of 40 mL (Comelli et al., 2008; Petterson et al., 2015). Samples were then agitated at 4°C on an orbital shaker set at 160 RPM for a period of 12 to 17 hours, then centrifuged at 10,000 x g for 45 minutes at 4°C. The supernatant was decanted, being careful to preserve any pellet. Samples were then centrifuged a second time at 10,000 x g for another 10 minutes and the remaining supernatant decanted. The resulting PCS and PGS pellets were transferred to a new RNase-free centrifuge tube and frozen at -80°C until RNA extraction.

### 2.4. RNA extraction

Viral RNA was extracted from PGS and PCS samples using the RNeasy PowerMicrobiome Kit (Qiagen, Germantown, MD), with the following deviations from the manufacturer’s recommended protocol: i) 200 mg of sample pellet was added to the initial extraction step in place of 200 μL of liquid sample, and ii) the optional phenol-chloroform mixture addition to the lysis buffer was substituted with Trizol LS reagent (ThermoFisher, Ottawa, Canada) to maximize lysis of cells/virion encapsulated fragments and protect RNA prior to vortexing and centrifugation. The resulting aqueous phase of the lysis procedure was then retained and processed as per the recommended protocol including the on-column enzymatic DNA removal step. RNA was eluted in 100 μl of RNAse-free water.

### 2.5. Viral recovery efficiency

An important metric in the quantification of viral signal in wastewater is the process recovery efficiency for targets of interest, as it facilitates a comparison of results from study to study, even if different sample processing or extraction methodologies/techniques are used. In this study, the efficiency of virus recovery following the fractionation, PEG concentration and RNA extraction process was determined by spiking vesicular stomatitis virus (VSV) and quantifying the recovered quantities of virus after sample processing. Spiking samples with a human coronavirus with low pathogenicity such as HCoV-229E as a recovery control (Gundy et al., 2009) was desirable but not practical due to the relative difficulty of its procurement in Canada at the time of this study and the difficulty of propagating coronaviruses in vitro. VSV is an enveloped, single stranded negative-sense RNA virus belonging to the *Rhabdoviridae* family, genus *Vesiculovirus* (Letchworth et al., 1999). The RNA genomes of both VSV and SARS-CoV-2 are encapsulated by a lipid envelope, and their particle sizes are similar; with VSV ranging from 70-200 nm (Cureton et al., 2010) and SARS-CoV-2 being approximately 100 nm (Supplemental Figure S2) (Bar-On et al., 2020). It was reasoned that these similar biophysical characteristics (lipid envelope and particle size) would lead both viruses to associate with wastewater matrices and to be precipitated with PEG with similar efficiencies. To maximize safety of the method, VSV was heat inactivated at 55°C for five minutes prior to use (Supplemental Figure S3).

Recovery efficiency was quantified twice during this study, following procedures similar to those outlined in Annex G of ISO 15216-1:2017 (ISO, 2017; Lowther et al., 2019; Randazzo et al., 2020). VSV was quantified via RT-ddPCR for both PGS and PCS from triplicate, serial dilutions of 5.5×10^4^, 5.5×10^5^, 5.5×10^6^ and 5.5×10^7^ copies VSV/μL of inactivated stock VSV culture spiked into the collected PGS samples and the PCS samples. Throughout the study, quantified quantities were not corrected for process extraction efficiency or for PCR inhibition. Three PCS and PGS samples were each spiked with 10 μL aliquots of 5.5×10^4^, 5.5×10^5^, 5.5×10^6^ and 5.5×10^7^ copies/μL. These samples were directly concentrated, extracted and quantified using RT-qPCR. The probes and primers used are listed in Supplemental Table S3. The VSV recovery efficiency (mean and standard deviation) was calculated based on the number of copies quantified using RT-qPCR. The equation for the calculations is as follows:

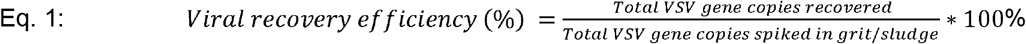

### 2.6. Variance of biomarkers for normalization

Analysis of variance was used to identify biomarkers with low variability and higher temporal consistency. The analysis of variance was conducted on 30 PGS and PCS samples over a period of 55 days (between April 8^th^ 2020 and June 2^nd^ 2020). The samples were analyzed for the following three internal normalization biomarkers: i) human microbiome-specific HF183 *Bacteroides* 16S ribosomal rRNA, ii) eukaryotic 18S rRNA and iii) PMMV.

### 2.7. RT-qPCR

Preliminary testing of samples with the CDC N1, N2 and N3 primer-probe sets and the Sarbeco E-gene primer-probe set (Supplemental Table S3) demonstrated best detection and least variance in technical replicates with the CDC N1 and N2 primer-probe sets. Singleplex, probe-based, one-step RT-qPCR (Reliance One-Step Multiplex RT-qPCR Supermix (Bio-Rad, Hercules, CA) was performed in this study using the 2019-nCoV Assay-RUO probe/primers mixes for CDC N1 and N2 gene regions (IDT, Kanata, Canada). All utilized primer/probe sets, their sequences and their sources (including PMMV and VSV) are described below in Supplemental Table 3. Reactions were comprised of 1.5 μl of RNA template input, 500 nM each of forward and reverse primers along with 125 nM of the probes in a final reaction volume of 10 μl. Samples were run in triplicate. Using a CFX Connect qPCR thermocycler (Bio-Rad, Hercules, CA), RT was performed at 50°C, 10 minutes, followed by polymerase activation at 95°C for 10 minutes, and 45 cycles of denaturation, annealing/extension at 95°C/10 s, then 60°C/30 s, respectively. Serial dilutions of the viral RNA standard were run on every 96-well PCR plate to produce standard curves used to quantify the copies of SARS-CoV-2 genes. In addition, RT-ddPCR-quantified pooled samples of RNA template were serial diluted and utilized to construct standard curves for PMMV normalization biomarker when RNA signal was normalized by the concentration of PMMV. Additionally, RT-qPCR runs were validated with the use of non-template-controls (NTCs), positive controls, negative controls of pre-COVID 19 pandemic wastewater samples and dilutions.

The limit of detection of the RT-qPCR assay was determined for N1 and N2 gene regions, by determining the number of copies per reaction which corresponds to a detection rate of ≥ 95% (<5% false negatives), as recommended by the MIQE guidelines (Bustin et al., 2009). Furthermore, samples were discarded if they did not meet the following conditions: i) standard curves with R > 0.95, ii) copies/reaction are in linear dynamic range of the curve and iii) primer efficiency between 90%-130%. Furthermore, sample replicates with values greater than 2 standard deviations from the mean of the triplicates were also identified as possible anomalies in this study and discarded.

### 2.8. RT-ddPCR

Singleplex, probe-based, one-step RT-ddPCR (1-Step RT-ddPCR Advanced Kit for Probes, BioRad, Hercules, CA) was used for absolute quantification of SARS-CoV-2 N RNA in wastewaters using the CDC N1, N2 or N3 primer-probe sets, or E RNA expression, using the Sarbeco E-gene primer-probe set (Supplemental Table S3). Primers and probes used in this study were obtained from Integrated DNA Technologies, Inc (IDT, Kanata, Canada) and ThermoFisher. 5 μl of RNA template, 900 nM each of forward and reverse primers and 250 nM of the probe together with the supermix were assembled in a final reaction volume of 20ul. Samples were prepared and run in triplicate. Droplet generation was performed using a QX200 droplet generator (Bio-Rad, Hercules, CA). Droplets were transferred to a new microplate, and PCR was completed in a C1000 (Bio-Rad, Hercules, CA) thermocycler as follows: reverse transcriptase (RT) was performed at 50°C, 60 minutes, followed by polymerase activation at 95°C for 10 minutes, and 40 cycles of denaturation, annealing/extension at 94°C/30 s, then 55°C/60 s, respectively. The polymerase was deactivated at 98°C for 10 minutes and droplets stabilized at 4°C for 30 minutes. Droplets were then read using a QX200 droplet reader (Bio-Rad, Hercules, CA). Positive droplets were called manually, and absolute quantification was performed using Quantasoft Analysis Pro v.1.0 (Bio-Rad, Hercules, CA). The limit of detection of the RT-ddPCR assay was determined for N1 and N2 gene regions by determining the number of copies per reaction which corresponds to a detection rate of ≥ 95% (<5% false negatives), as recommended by the MIQE guidelines (Bustin et al., 2009).

### 2.9. Statistical analysis

In order to test for significant differences between data sets comparing the detection of SARS-CoV-2 in PGS and PCS samples, chi-square and Fisher’s exact test statistical analyses were conducted using GraphPad’s Prism 8.3 software (La Jolla, CA). A student’s t-test was used to test for statistical differences between detection of RNA in RT-qPCR and RT-ddPCR assays for PGS and PCS. A student’s t-test and Pearson’s correlation analyses were performed to test for significance and the strength of the correlation between RNA signal and epidemiological data, with a *p*-value of 0.05 or lower signifying significance.

## 3. Results & discussion

### 3.1. Viral RNA recovery efficiency

The recovery through the concentration and extraction steps was quantified by spiking samples with serial dilutions of inactivated VSV. The percent recoveries for VSV spiked in PGS and PCS were 8.4 ± 3.6% and 9.3 ± 4.9%, respectively. The recovery of the surrogate virus through both PCS and PGS concentration and extraction was similar with all spiked-in quantities. Other recent studies investigating surrogate virus recoveries following similar PEG concentration reported variable results for various surrogates; <6% recovery of murine hepatitis virus (MHV) (Ye et al., 2016) along with reported recoveries of 33.3 ± 15.6% and 57% of *Escherichia virus* MS2 (MS2) by Balboa et al. (2020) and Kumar et al. (2020). Other concentration methods have also been used, such as ultrafiltration and ultracentrifugation (~20% to 33.5% recovery efficiency of MHV) (Ahmed et al., 2020b; Ye et al., 2016) and aluminum hydroxide adsorption-precipitation (30.4 ± 11.0% recovery of *Mengovirus* (MGV)) (Medema et al., 2020). It is important to recognize that each study used slightly different methods and viral surrogates, making it difficult to make direct comparisons and generalizations (Lu et al., 2020; Michael-Kordatou et al., 2020). Each surrogate virus will differ in how it interacts with wastewater and this may also be dependent on the characteristics of the wastewater as well as the properties of the virus/fragment that may have very different partitioning/degradation characteristics. It is unclear yet how effective filtration-based concentration techniques perform with high-solid samples, especially with viruses that are highly associated with solids. When analyzing high solids containing samples, such as PGS and PCS, PEG precipitation or other flocculation approaches may be more effective due to an incompatibility of this matrix with ultrafiltration due to possible complication associated with membrane clogging. The advantages of using PGS and PCS, which may have a greater and more consistent RNA signal, should be balanced against the apparent lower recovery of PEG precipitation. Additional studies are needed to develop and assess appropriate and effective methods and surrogates for analysis of SARS-Cov-2 in wastewaters.

### 3.2. Comparison of RT-qPCR and RT-ddPCR for the detection and quantification of SARS-CoV-2 RNA

This study tested the detection and quantification of RT-ddPCR and RT-qPCR for SARS-CoV-2 RNA signal in PGS and PCS samples. The in vitro transcribed RNA was observed to be reliably detected with primer-probe RT-ddPCR assays to a limit of detection of 5 copies/reaction in both N1 and N2 RT-ddPCR assays. This is consistent with the purported high sensitivity of the digital PCR technology. In vitro transcribed viral RNA was detected to a limit of detection of 2 copies/reaction in both the N1 and N2 RT-qPCR assays, (using the high sensitivity Bio-Rad One-Step Reliance Supermix (Bio-Rad, Hercules, CA)), which was unexpected when comparing to the RT-ddPCR limit of detection. The standard curves utilized for the quantification of different RNA targets for RT-qPCR are as follows: N1 (slope: -3.372, intercept: 38.184, R^2^: 0.972, E: 97.96%), N2 (slope: -3.179, intercept: 37.870, R^2^: 0.954, E: 106.32%), PMMV (slope: -2.806, intercept: 39.142, R^2^: 0.968, E: 127.17%) and VSV (slope: -3.518, intercept: 39.846, R^2^: 0.995, E: 92.41%). The standard curves demonstrate good linearity for RT-qPCR in a range between 2 to 60 copies/reaction for N1 and N2, 1.4 × 10^2^ to 3.6 × 10^4^ copies/reaction for PMMV and 1.6 × 10^0^ to 1.6 × 10^4^ copies/reaction for VSV.

A comparison was performed between the one-step RT-qPCR and RT-ddPCR using the same singleplex N1 probe-primer set for the quantification of SARS-CoV-2 in solids-rich, low concentration SARS-CoV-2 RNA signal wastewaters (Figure 3). Six PGS samples and five PCS samples were analyzed using RT-qPCR and RT-ddPCR. All samples were collected from the two cities during the same low incidence periods (<60 active cases / 100,000 people) case number study period allowing the assessment of quantification and degree of variability in samples with low RNA concentrations. The mean and standard error of the PGS samples analyzed during the same period of low incidence cases are 133.4 ± 9.0 and 167.1 ± 25.6 N1 gene copies/100 μL of extracted RNA for RT-ddPCR and RT-qPCR, respectively. Meanwhile, the mean and standard error of the PCS samples are 33.5 ± 5.8 and 130.4 ± 20.8 gene copies/100 μL of extracted RNA for RT-ddPCR and RT-qPCR, respectively (Figure 3). Although a significant decrease in detected copies for PCS samples with RT-ddPCR is observed, it is noted that the coefficient of variation (%CV) for the ddPCR assay (38.4%) compared to qPCR (35.7%). While the %CV for PGS samples for the ddPCR assay is lower (16.5%) compared to qPCR (37.5%).

**Figure 3:**
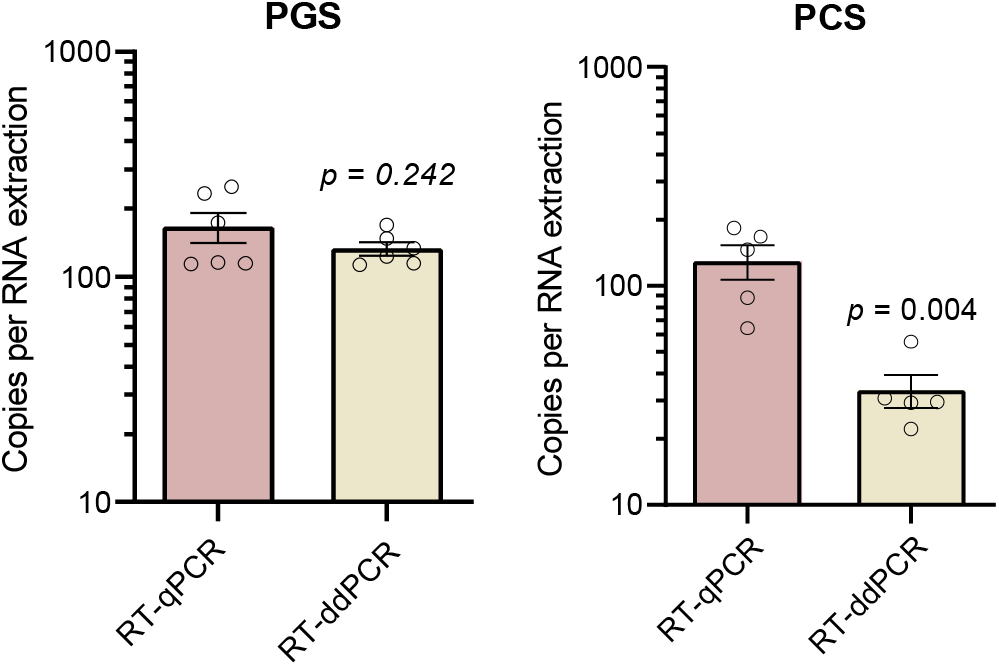
Comparison of copies per 100 pL of extracted RNA in RT-qPCR and RT-ddPCR for PGS (n=6) and PCS (n=5).

The difference in quantification of the PCS samples between the N1 RT-qPCR and RT-ddPCR assays suggests inhibition of the reverse transcription and/or polymerase chain reaction of the PCS sample. Given that this assay partitions the sample volume into approximately 1 nL droplets, it’s conceivable that the effective concentration of any RT and/or PCR inhibitors present in the PCS matrix are markedly increased. In contrast, RT-qPCR is performed in a non-partitioned assay volume and may thus be less sensitive to inhibition. The apparent inhibition in ddPCR may also be explained by differences in the reagents used for the RT-ddPCR and RT-qPCR assays. The inhibition resistance/inhibitor removal of the high sensitivity RT-qPCR reagent appears to provide better detection when utilized in pegged sludge matrices. Quantification of two-fold and five-fold dilutions of PCS samples was performed and support the theory that RT-ddPCR was likely inhibited; RT-qPCR shows good quantification of diluted samples while RT-ddPCR suffered from inhibition. These findings contradict the theoretical assumption that RT-ddPCR is less prone to inhibition due to relative insensitivity to differences in amplification efficiencies (due to its binary “all-or-nothing” reporting of amplification) (Salipante and Jerome, 2020). However, at least one report found that undiluted raw wastewater inhibits one-step RT-ddPCR amplification of PMMV RNA to the same degree as the RT-qPCR assay (Rački et al., 2014). Given that RNA in both PGS and PCS samples was at a very low concentration, approaching the limits of detection, it is highly likely that inhibitors in the PCS matrix are responsible for the decreased sensitivity observed in RT-qPCR vs. RT-ddPCR.

Of note, it was also attempted in this study to use a commercially available multiplex RT-ddPCR assay that employs primer-probe sets amplifying N1, N2 and N3 regions of the viral N RNA as well as a human transcript (SARS-CoV-2, Bio-Rad). However, it was determined that the discrimination between positive and negative droplets (fluorescence amplitude) was poor, making quantitative analysis impossible. RT-ddPCR has a myriad of theoretical advantages such as absolute quantification that is not dependent on calibration curves, insensitivity to common PCR inhibitors, and the ability to multiplex (Salipante and Jerome, 2020). There is a need to explore this further in future studies and to optimize these methods and quantification techniques for wastewater samples. However, based on the better detection using the current RT-qPCR approach, this method was utilized for the remainder of this study to quantify SARS-CoV-2 in both PGS and PCS solids from the Ottawa and Gatineau WRRFs.

### 3.3. Detection and variance of SARS-CoV-2 viral RNA in PGS and PCS

In this study, the sensitivity and variability of the RT-qPCR assay in PGS and PCS was compared by investigating the percentage of sample replicates; with replicates including repeated RNA extraction step and PCR quantification samples along with technical triplicates. The limit of detection used in the study for RT-qPCR assays is described above. Replicate runs (24 paired PGS and PCS samples, for a total of 72 technical replicates each) were collected on the same dates across 83 days. PCS samples collected and analyzed at the same time as PGS samples over a 3-month period exhibited stronger percent detection for N1 (92.7% for PCS compared to 79.2% for PGS, *p* = 0.007) and N2 (90.6% for PCS compared to 82.3% for PGS, *p* = 0.092) (Figure 4). Variance in percent detection of PCS was shown to be similar for all samples, with coefficients of variation ranging from 29.1% to 31.7%.

**Figure 4:**
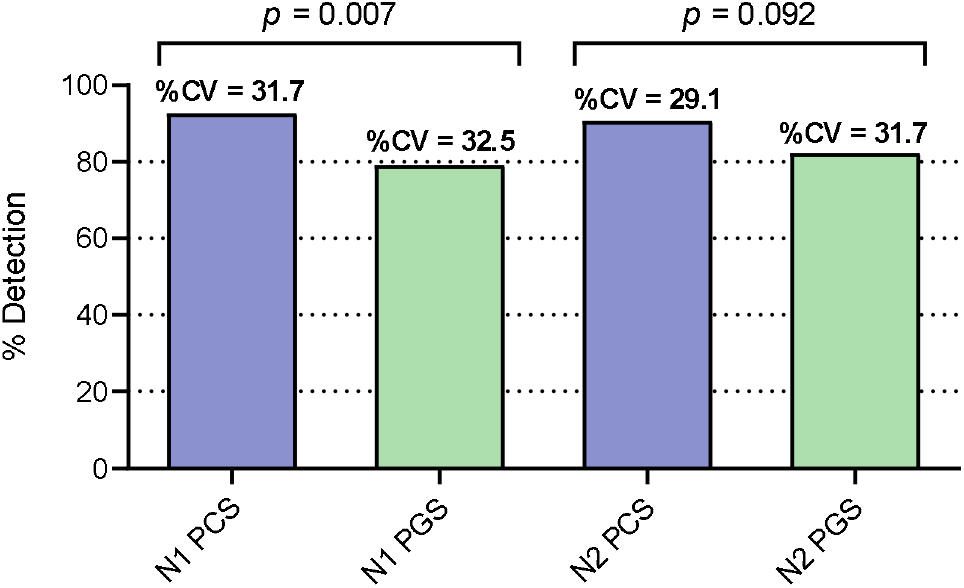
Sensitivity of N1 and N2 RT-qPCR assays comparison between PCS and PGS samples. Significance between detections established using Chi-Squared test. Variance is shown with %CV (n=24).

The decreased sensitivity of SARS-CoV-2 detection in PGS samples could be due to the increased susceptibility of solid particulate matter in this wastewater fraction to daily fluctuations in flowrate and wastewater biochemical characteristics at the WRRFs compared to the sludge samples collected in the primary clarifier stream. In addition, the PGS samples undergo a laboratory settling step in this study to isolate the settled solids from the liquid fraction of the sample. This additional step (which is not applied to the PCS samples) may also contributes to the lower percent detection of SARS-CoV-2 signal of these samples due to increased holding times and processing times. As such, the result of this study confirms PCS samples as the high-solids samples that demonstrate an elevated frequency of detection of SARS-CoV-2 N1 and N2 RNA in municipal wastewaters during decreasing and low incidence of community COVID-19 (Alpaslan-Kocamemi et al., 2020; Balboa et al., 2020; Peccia et al., 2020a). In addition, it is noted that the viral RNA longitudinal trendline from the PGS samples did not show strong correlation with either the trendline from the PCS samples, or municipal epidemiological data, further supporting PCS sampling as the more robust basis for community COVID-19 monitoring in wastewater solids.

### 3.4. Variability of normalization biomarkers

A multitude of systematic variations exist in molecular wastewater surveillance that makes it challenging to accurately measure SARS-CoV-2 RNA across days, months and years. These include, but are not limited to: diurnal variation in plant flow, changes in gross proportions of solids, sample collection and storage, sample processing and sample analysis. Due to these factors, a critical aspect of wastewater epidemiology is sample normalization (Armanious et al., 2016). The necessity to normalize SARS-CoV-2 RNA data has also been identified in more recent studies (Alpaslan-Kocamemi et al., 2020; Balboa et al., 2020; Kaplan et al., 2020; Peccia et al., 2020a; Wu et al., 2020). To compare the variability and temporal consistency of biomarkers in this study 8 PCS samples (24 including technical triplicates) were analyzed using RT-qPCR for all three biomarker gene regions: human-specific HF183 *Bacteroides* 16S rRNA, human eukaryotic 18S rRNA and PMMV. All three tested biomarkers were detected in PCS samples with a relatively high level of incidence. While all three RNA targets were detected in PCS samples, it was observed that the distribution of their expression (i.e. quantified through an analysis of variance) of the fecal biomarker PMMV was lower as compared to the 16S and 18S biomarkers in PCS samples (Figure a). The lower variability of PMMV (C_t_ variance = 1.18) compared to 16S (C_t_ variance = 5.32) and 18S (C_t_ variance = 5.12) may be due to the relative toughness and stability of the virus in difficult environments (Kitajima et al., 2018). Furthermore, the viral fragments of this biomarker may preferentially adhere to the solids fraction of wastewaters via electrostatic and/or hydrophobic effects (Armanious et al., 2016). Additionally, in order to quantify the variance of the normalization biomarkers in this study, the samples run this comparison were also verified across each surveyed WRRF independently (Figure 5b). The PMMV internal normalization biomarker shows an improved consistency and lower variability (maximum change in C_t_; ΔC_t_ = 0.01) between the WRRFs compared to 16S (ΔC_t_ = 1.47) and 18S (ΔC_t_ = 2.30); which demonstrates a relative steady signal between differing WRRFs. Due to the consistency of the PMMV fecal biomarker in the PCS samples across 55 days of sampling, PMMV was utilized in this study as a SARS-CoV-2 N1 and N2 RNA internal control for PCS samples. The low variance of PMMV in PCS, coupled with the use of PMMV as an internal normalization biomarker, was also recently reported by Wu et al. (2020).

**Figure 5:**
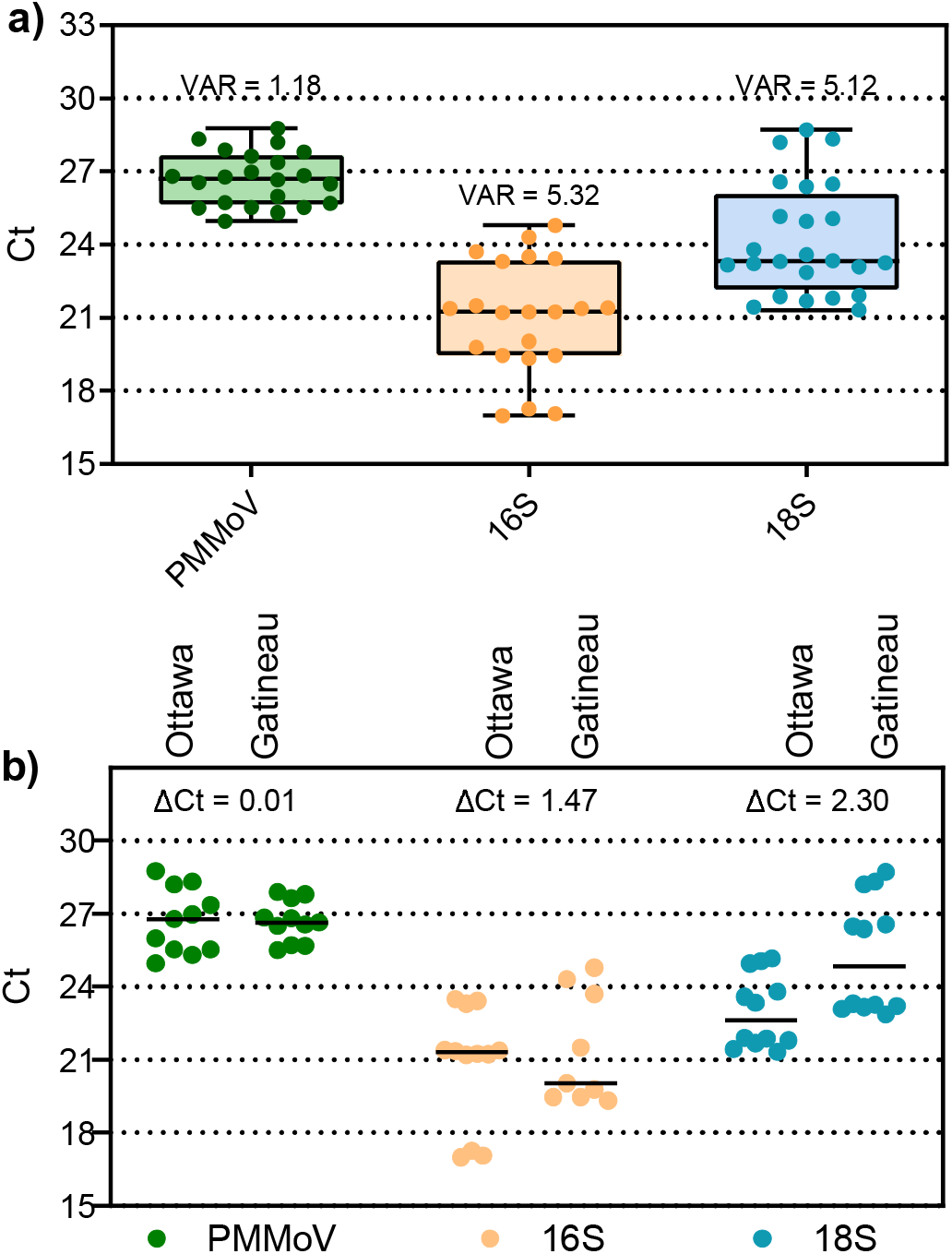
Variance of PCS normalization biomarkers, a) combined data set comprised of both cities samples, and b) data set separated by city. Analysis of variance and maximum change in Ct (ΔCt) (n=8).

### 3.5. Quantification of SARS-CoV-2 viral RNA in PCS and correlation with COVID-19 case data

As PCS was identified as the solids-rich sample showing the highest RNA detection rate, SARS-CoV-2 RNA was measured in PCS samples from Ottawa and Gatineau WRRFs between April 1st and June 30th, 2020. This sampling period encompasses a decreasing COVID-19 prevalence in two cities (peaks of 56.7 and 57.3 confirmed cases/100K inhabitants in Ottawa and Gatineau respectively) as well as an ensuing period of low prevalence characterized by many days with low new daily reported cases (56.7 → 4.8 and 57.3 → 10.2 confirmed cases/100K inhabitants in Ottawa and Gatineau, respectively). In addition to the technical triplicates of each sample, five of the 14 samples in Ottawa were re-extracted and re-quantified via RT-qPCR. In Gatineau, four of the 8 samples were re-extracted and re-quantified via RT-qPCR. Two distinct but complementary normalization approaches were applied to the observed SARS-CoV-2 RNA signal to account for variations in WRRF wastewater flow, composition and treatment along with temperature, time variations in travel and storage along with human errors in the processing of the samples. In particular, this study normalizes the RNA signal for i) the WRRF mass flux of solids in the sampled primary clarifier stream and ii) the PMMV internal normalization biomarker expression. The SARS-CoV-2 viral RNA copies/L and the normalized viral data are benchmarked against and correlated to epidemiological metrics provided by the Ottawa public health agency and the regional public health agency of the city of Gatineau.

Three epidemiological data sets based on clinical testing were identified by the local public health agencies as best estimates of COVID-19 prevalence in the two cities: i) daily new cases of COVID-19, ii) active cases of COVID-19 based on an active period of fourteen days, and iii) percent positive of total daily reported clinical COVID-19 tests performed. Two key factors/limitations are noted with respect to these epidemiological data sets shown in this study. Firstly, the testing at the onset of the pandemic (March and April 2020) was variable and low in both cities due to limitations in human resources, laboratory reagents and testing equipment. Hence, the first four weeks of the twelve-week period for which wastewater samples were profiled were subject to variable and lower testing rates per day that likely under-reported both the number of new cases and active cases during this period. Secondly, early testing/screening was less available to the general population in both cities, with testing heavily biased towards hospitalized patients and health care workers. This potentially artificially inflates the percent positive data during the first four weeks of the study, with the effect on the percent positive being likely lesser than the effect of limited testing on the total case numbers.

### SARS-CoV-2 RNA in PCS and correlation with COVID-19 case data

The average and standard deviation of technical triplicates and extraction replicates that repeated the concentration, extraction and RT-qPCR steps (shown as error bars in Figures 6 and 7) for the longitudinal viral RNA data sets in this study are plotted along with a percent positive and seven day floating average percent positive epidemiological data sets. Due to limited testing during the first four weeks of the longitudinal study, percent positive was identified as a potentially useful epidemiological metric of COVID-19 prevalence to compare to SARS-CoV-2 RNA measurements in wastewater. As such, this metric is included in Figures 6 and 7 to be benchmarked against the measured SARS-CoV-2 signal.

N1 and N2 RNA signal is first expressed in copies/L (of PCS) in this study (Figures 6a and 7a). Equivalent volumes of PCS were PEG concentrated and RNA extracted throughout the sampling period. As expected, and similar to other studies investigating primary sludge and wastewater solids, the raw copies/L data sets for the two cities (Figures 6a and 7a) are relatively noisy with no clear trend observed (Medema et al., 2020; Randazzo et al., 2020; Wu et al., 2020). The observed concentrations in this study (1.7 × 10^3^ to 7.8 × 10^4^ copies//L (Ottawa) and 6.6 × 10^4^ to 3.8 × 10^5^ copies/L (Gatineau)) are in agreement with other studies investigating SARS-CoV-2 RNA viral signal in PCS. Concentration ranges of 1.7 × 10^6^ to 4.6 × 10^8^ copies/L, 1 × 10^4^ to 4 × 10^4^ copies/L and 1.2 × 10^4^ to 4.0 × 10^4^ copies/L have been reported in PCS by Alpaslan-Kocamemi et al., (2020), Balboa et al., (2020) and Peccia et al., (2020b), respectively.

Although the N1 and N2 RNA genes show similar longitudinal trends to each other in both the Ottawa and Gatineau WRRFs (Figures 6a and 7a), the inherent variations in signal results in noise, making it difficult to identify real changes in viral signal. In particular, this is seen in the large amplitudes of the standard deviations of many data points in the longitudinal data sets of Ottawa and Gatineau. The noise in the RNA data may be caused by inherent, weather-induced random variations in wastewater biochemical characteristics, solid composition and flowrate (e.g., due to weather, changes in daily household water consumption, etc.) as well as potentially significant effects associated with the collection and transport of the samples and RNA concentration, extraction and analysis. The copies/L longitudinal data in Ottawa and Gatineau clearly demonstrate that SARS-CoV-2 quantification in wastewater is inherently noisy and hence normalization of the data should be explored.

No significant correlation between N1 and N2 at either the Ottawa WRRF or the Gatineau WRRF is observed across the study time period (Table 2). Strong and significant correlation would have suggested that SARS-CoV-2 RNA might be intact in PCS prior to concentration and extraction, which is not herein observed in this study. Critically, when comparing either N1 or N2 copies/L to each of the epidemiological metrics (daily cases, active cases and percent positive) it appears that in Ottawa no correlation exists between the N1 or N2 RNA copies/L signal and any of the three epidemiological metrics. Meanwhile, in Gatineau, significant correlations exist between the N1 and N2 copies/L signal and epidemiological data sets, with the strongest correlations being observed with the number of active cases (Table 2).

### Mass flux of primary clarified sludge copies SARS-CoV-2 RNA per day and correlation with COVID-19 case data

To correct for systematic variability, the first normalization approach applied in this study is to normalize the N1 and N2 RNA signal to both the mass of the PEG-concentrated solids subject to nucleic acid extraction and also the daily mass flux (mass of volatile solids (VS) solids through the primary clarifier stream per day) at each WRRF (Figures 6b and 7b). This normalization approach results in units of N1 and N2 copies/day as a solids mass flux basis through the WRRF. This normalization approach is intended to compensate for variations in solids concentration and flowrate in the primary clarifier stream at the WRRF due to weather effects, precipitation and infiltration/inflow in the sewers.

When comparing longitudinal plots in Ottawa of copies/L (Figure 6a) to copies/d (Figure 6b), the variance of the solids mass flux normalized data set of copies/d does not appear to have been significantly reduced the systemic noise of the copies/L data sets. Similar findings are observed for the Gatineau normalized data (Figures 7a and 7b). The substantial noise maintained in the solids mass flux normalized data sets of the two cities and the significantly large standard deviations of longitudinal data points indicates that the fluctuations associated with the solids concentration and flowrate in the primary clarifier stream was likely not a dominant source of the inherent variance in the copies/L data sets.

As observed for the copies/L data, the correlation between the N1 and N2 data sets were not significant for either Ottawa or Gatineau. Further, the normalization of the N1 and N2 data for solids mass flux at the WRRFs appear to worsen correlations, with anticorrelations increasing, for all three epidemiological metrics in Ottawa and Gatineau (Table 2). This lack of impact when normalizing operational mass flux of solids at the two WRRFs in this study is likely due to the fact that both WRRFs directly control the flow of the primary clarifier stream at their respective facilities, hence reducing the variation in the flux of solids and in turn minimizing the impact of this variation in WRRF operation on the N1 and N2 signal. Thus, systematic variation in the data sets are likely associated with the sample collection and storage along with RNA concentration, extraction and analysis steps performed in the study.

### PMMV-normalized SARS-CoV-2 RNA in PCS and correlation with COVID-19 case data

PMMV is the most abundant human fecal RNA virus (Kitajima et al., 2018) and has been previously proposed as a biomarker for fecal contamination in water (Hamza et al., 2011; Rosario et al., 2009). PMMV has also more recently been used as an internal reference for SARS-CoV-2 in wastewater (Wu et al., 2020). The second normalization approach applied in this study is the division of the RNA N1 and N2 copies by PMMV copies. Due to its low variability and high expression in PCS, PMMV was identified as the preferred internal reference of the three tested biomarkers tested in this study.

PMMV normalization appears to sufficiently reduce background noise associated with systematic variations in the Ottawa and Gatineau WRRF RNA signals that are possibly associated with the collection and transport of the samples along with the RNA concentration, extraction and analysis steps of PCS RNA signal during decreasing and low incidence periods of COVID-19 disease in this study (Figure 6c and 7c). In particular, the amplitude of the standard deviation associated with each data point in the longitudinal data sets of Ottawa and Gatineau decreased. This increase in precision ultimately allows for greater distinction between low-incidence data points, hence enabling improved identification of trends in the data sets.

Correlation between the PMMV normalized N1 and N2 signals remained insignificant in Ottawa and Gatineau (Table 2). However, this normalization approach also outlines strong, significant and positive correlations between both the N1 gene and the N2 gene with all three epidemiological data sets in Ottawa. The strongest correlation between N1 and N2 PMMV normalized RNA signal is observed with the 7-day rolling average percent positive epidemiological metric in Ottawa. Although the percent positive data during the first four weeks of the study may be biased towards hospitalized patients and health care worker testing, this clinical testing metric in Ottawa is identified as the preferred metric by the public health unit of the city (for the reasons described above). As such, decreasing the systematic variation in the data sets via PMMV normalization establishes a modified trend of the RNA signal and this trend shows the strongest correlation of the RNA signal to city’s identified preferred epidemiological metric.

Strong, significant and positive correlation is also shown between the N1 PMMV normalized RNA signal with the active cases epidemiologic metric in Gatineau; while the N2 PMMV normalized signal shows moderate, significant correlation to the active cases. Although the PMMV normalized Gatineau RNA signal data shows agreement with the active cases epidemiological metric, results were varied when correlated to daily cases and 7-day rolling average percent positive. The strongest correlation observed in this study for the Gatineau RNA signal and the epidemiological metrics of the City exists between both the N1 and N2 copies/L RNA signal and the active cases. It is also noted that the longitudinal trends in N1 and N2 PMMV signals in Gatineau are similar to those in Ottawa. This is expected as the two cities are geographically close with many inhabitants travelling across bridges between the cities. The observed differences in correlations between SARS-CoV-2 RNA signal in wastewater to clinical testing metrics in the two neighboring cities of this study is illustrative of the challenges associated with interpreting and correlating RNA signal acquired from distinct WRRFs to clinical testing metrics acquired from distinct health agencies.

**Figure 6:**
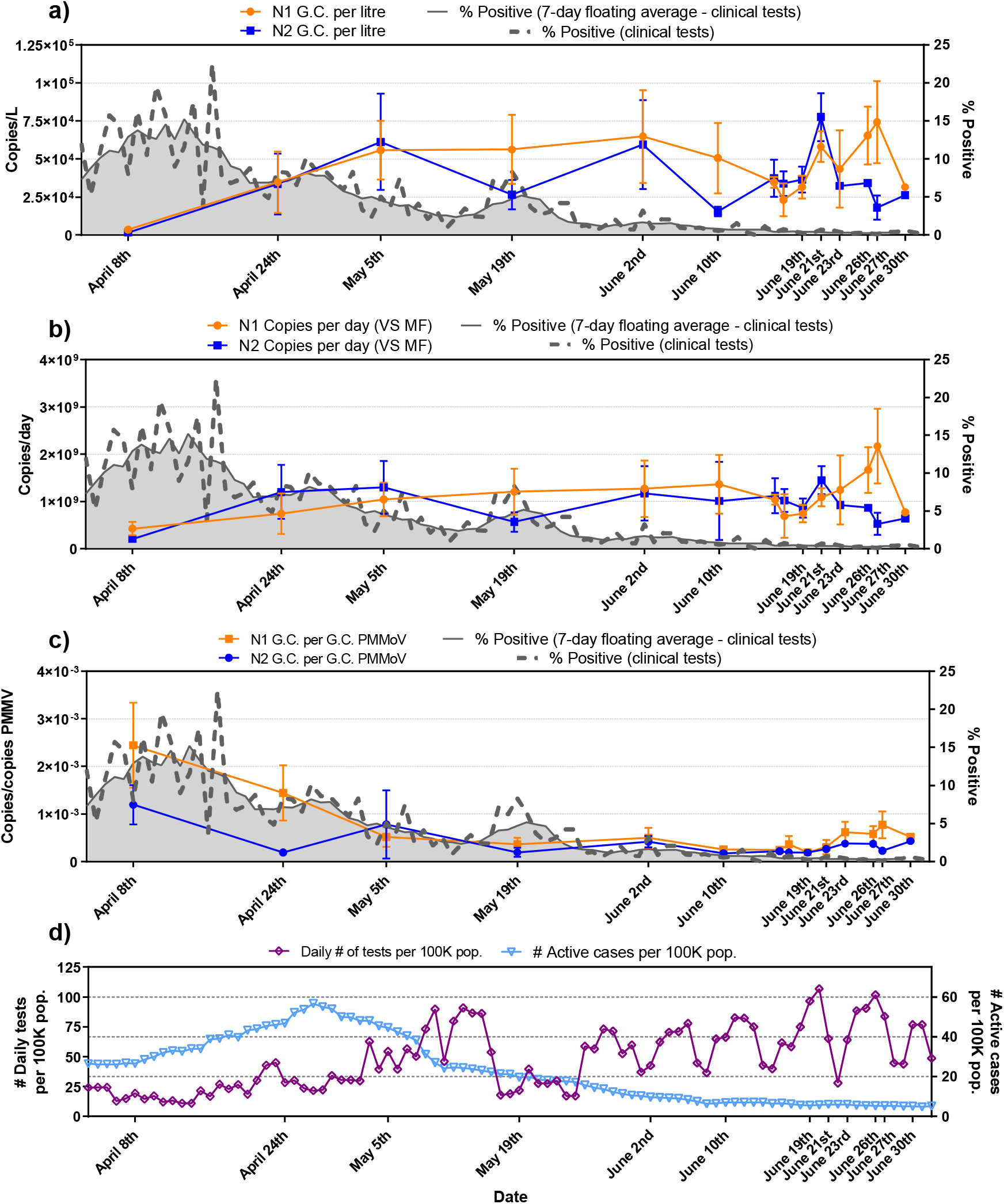
Trends of N1 and N2 SARS-CoV-2 viral copies with epidemiological metrics, a) copies/L of PCS, b) copies/d that was normalized by the mass flux through primary clarifier and c) copies/copies of PMMV that was normalized by PMMV.

**Figure 7:**
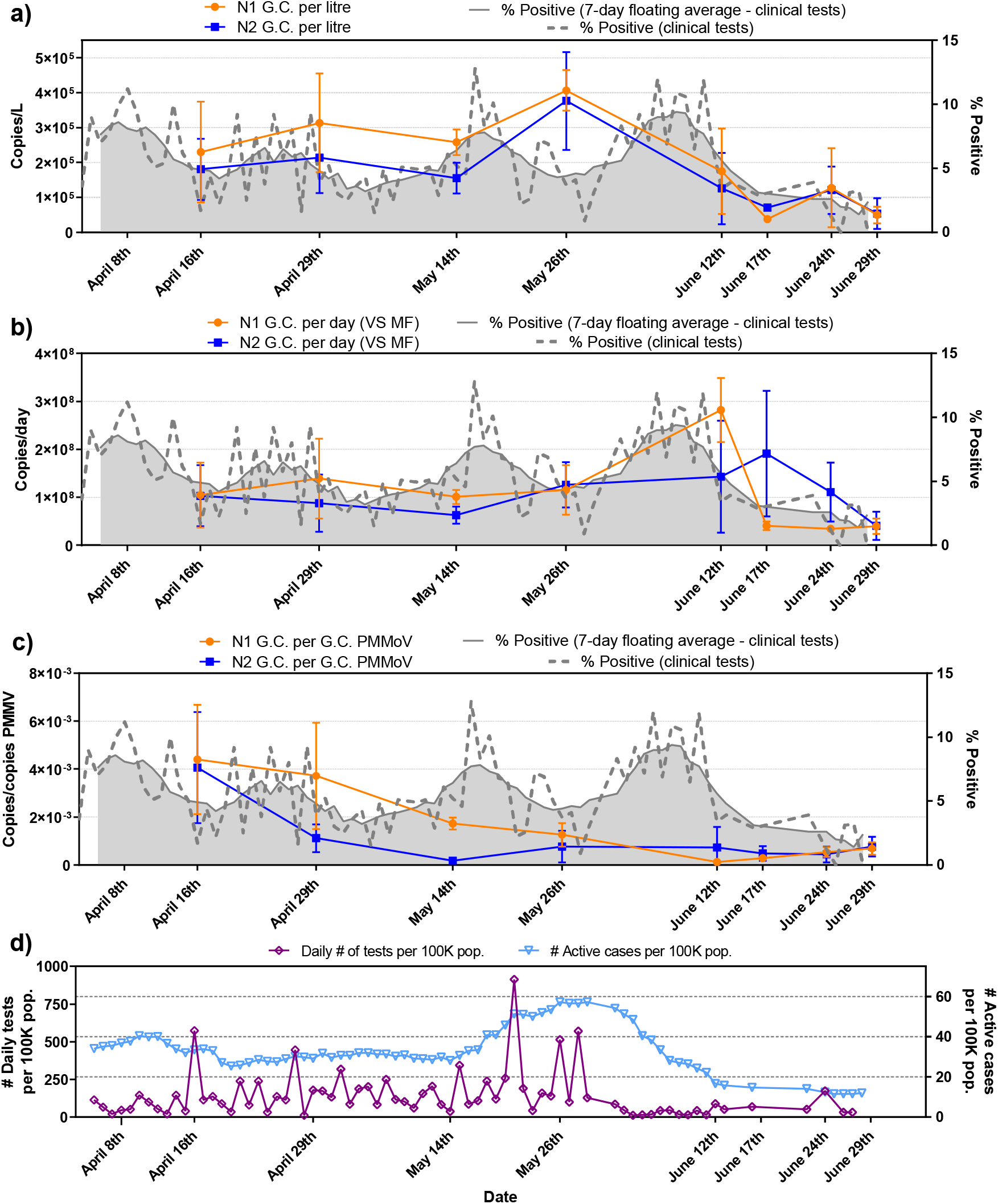
Trends of N1 and N2 SARS-CoV-2 viral copies with epidemiological metrics, a) copies/L of PCS, b) copies/d that was normalized by the mass flux through primary clarifier and c) copies/copies of PMMV that was normalized by PMMV.

**Table 2:**
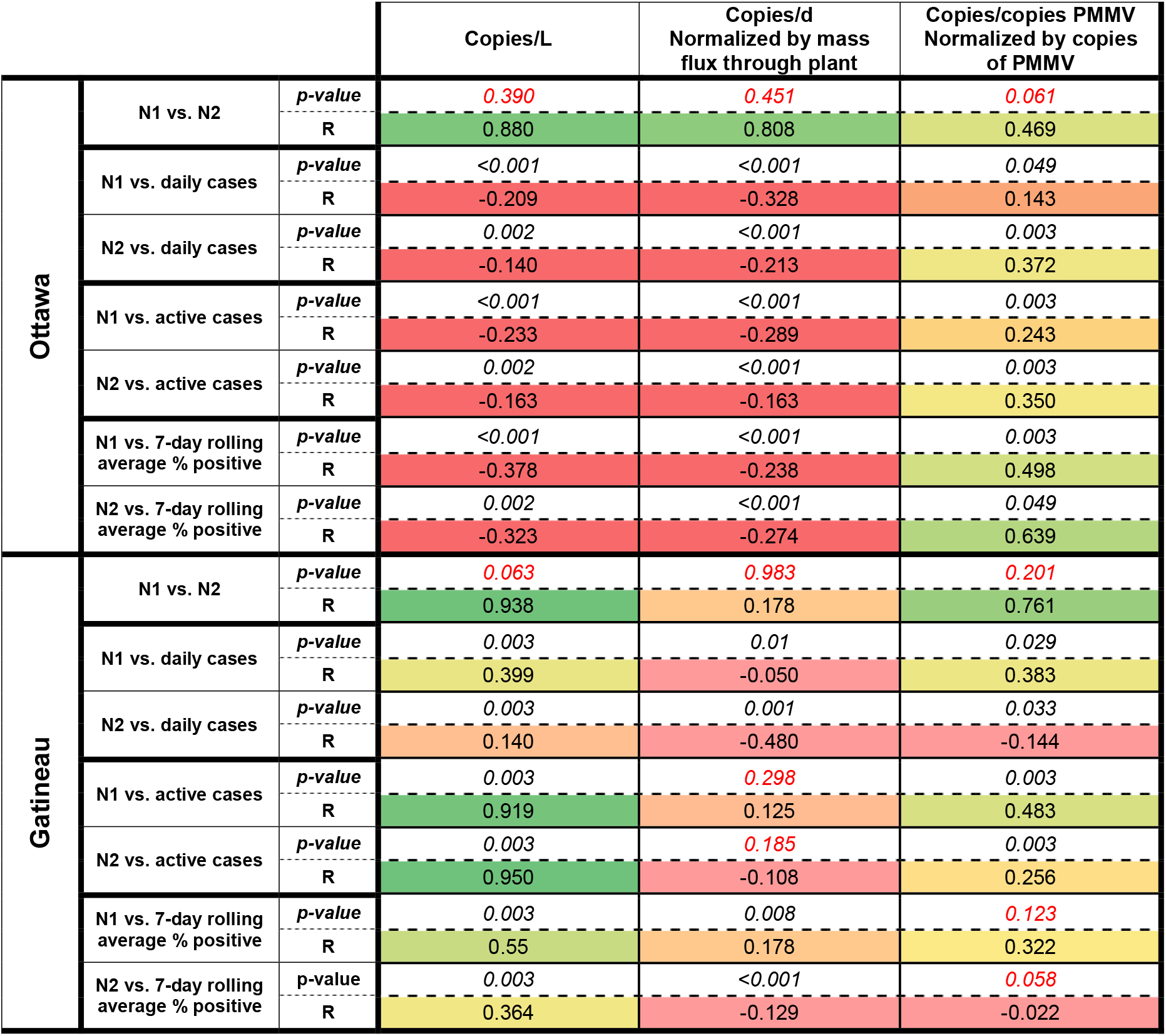
Correlation analyses between SARS-CoV-2 RNA signal in PCS in the Ottawa and Gatineau WRRFs with epidemiological metrics. RNA signal is expressed as copies/L of PCS, copies/d that was normalized by the mass flux through primary clarifier, and copies/copies of PMMV that was normalized by PMMV.

## 4. Conclusion

This study is the first investigation and detection of SARS-CoV-2 trends in wastewater in Canada. It identifies primary clarified sludge as a preferred solids-rich sample compared to post grit solids for the detection of SARS-CoV-2 signal during decreasing and low incidence of viral load in communities. Based on the reagents used in this study, RT-qPCR shows superior quantification of SARS-CoV-2 N1 and N2 gene signal in primary clarified sludge compared to RT-ddPCR. Finally, it is demonstrated that PMMV is a potential effective normalization biomarker for RNA signal to reduce noise inherent to the WRRF operation along with the sampling, transport and processing of the samples. The normalization of N1 and N2 SARS-CoV-2 signal using PMMV enables strong correlation to epidemiological metrics in two surveyed WRRFs across decreasing and low-incidence cases of COVID-19.

## Data Availability

Data is available upon request.

## Acknowledgements

The authors wish to acknowledge the help and assistance of the Dr. Marc-André Langlois of the University of Ottawa, the Children’s Hospital of Eastern Ontario’s Research Institute, Ms. Tammy Rose, Mr. Pawel Szulc and Mr. Tyler Hicks the City of Ottawa, Mr. Fabien Hollard of the City of Gatineau, Dr. Monir Taha of Ottawa Public Health, Mr. François Tessier of le Centre intégré de santé et de services sociaux de l’Outaouais (CISSSO), Public Health Ontario and l’Institut national de santé publique [Québec] (INSPQ) and all their employees involved in the project during this study. Their time, facilities, resources and assistance provided throughout the study greatly contributed to this work.

## Supplemental Material

**Figure S1:**
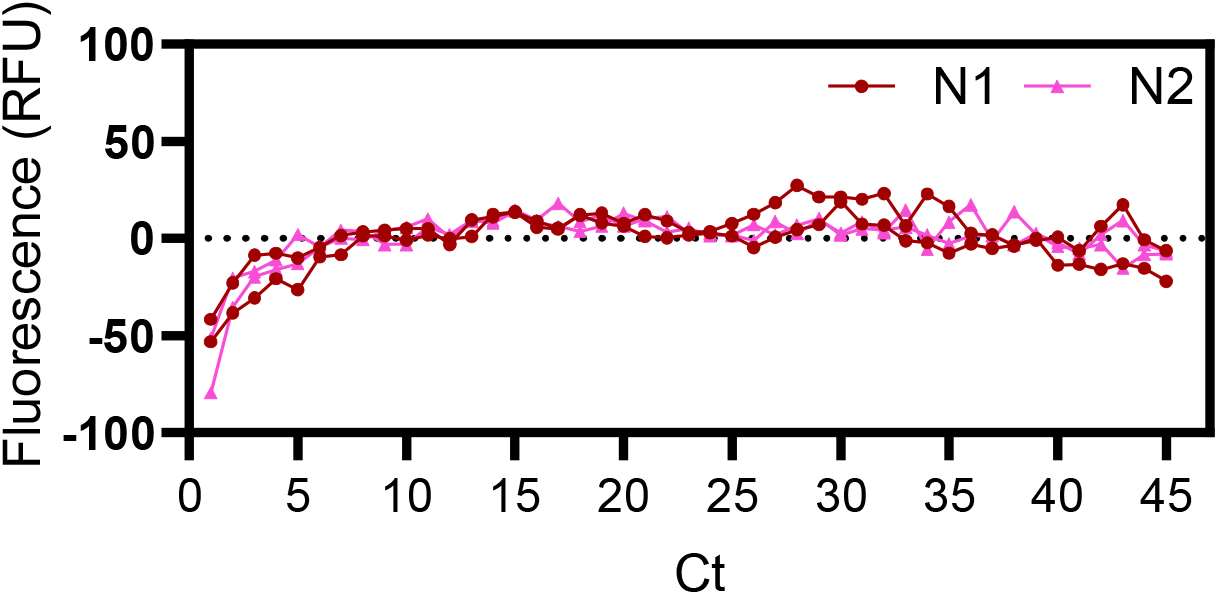
Amplification curves confirming that the bio-banked sample from Aug. 2019 (prior to pandemic) is a negative sample, quantified for the N1 and N2 SARS-CoV-2 genes.

**Figure S2:**
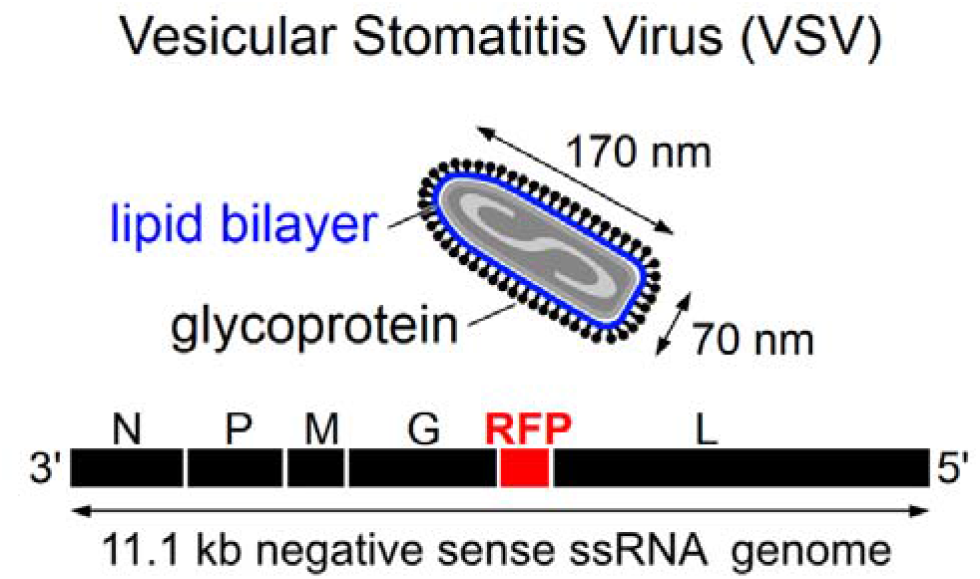
Schematic of VSV, lipid bilayer and glycoprotein identified.

**Figure S3:**
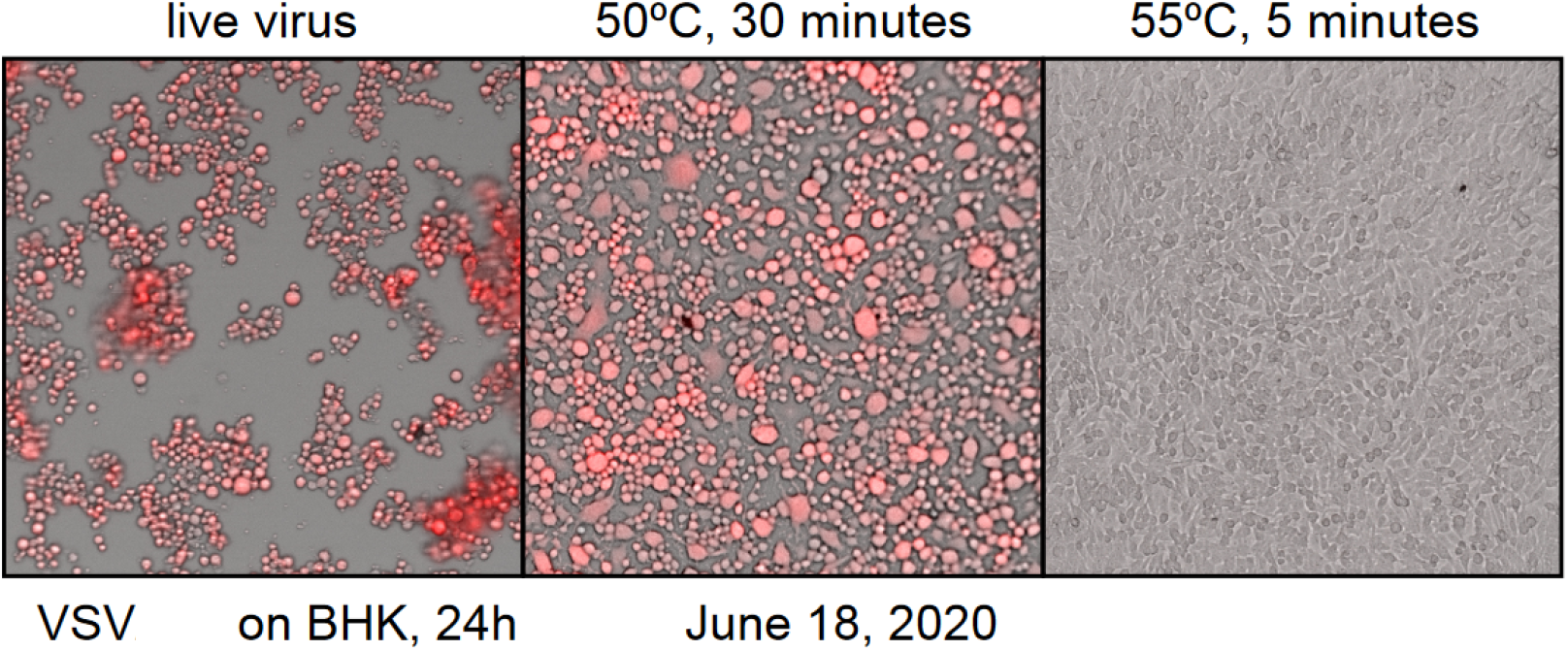
Confirmation of VSV heat inactivation.

**Table S1:** Average and standard deviations of wastewater quality characteristics of the PGS samples across the study period.

| Ottawa | PGS |  | Temp.<br>(°C) | pH | DO<br>(mg/L) | sBOD<br>(mg/L) | sCOD<br>(mg/L) | TAN<br>(mg-N/L) | Turbidity<br>(NTU) | TSS<br>(mg/L) | VSS<br>(mg/L) |
| --- | --- | --- | --- | --- | --- | --- | --- | --- | --- | --- | --- |
|  |  | Average | 16.75 | 7.41 | 4.05 | 7.81 | 70.04 | 19.22 | 129.79 | 162.39 | 153.83 |
|  |  | Standard deviation | 3.47 | 0.23 | 1.48 | 5.32 | 15.55 | 16.20 | 44.28 | 73.12 | 70.95 |
| Gatineau | PGS |  | Temp.<br>(°C) | pH | DO<br>(mg/L) | sBOD<br>(mg/L) | sCOD<br>(mg/L) | TAN<br>(mg-N/L) | Turbidity<br>(NTU) | TSS<br>(mg/L) | VSS<br>(mg/L) |
|  |  | Average | 15.46 | 7.43 | 4.96 | 7.60 | 73.38 | 12.19 | 128.22 | 144.77 | 129.88 |
|  |  | Standard deviation | 4.96 | 0.25 | 2.05 | 3.90 | 23.89 | 10.99 | 99.34 | 125.80 | 113.64 |

**Table S2:** Average and standard deviations of wastewater quality characteristics of PCS samples across the study period.

| Ottawa | PCS |  | TS (g/L) | VS (g/L) | % VS |
| --- | --- | --- | --- | --- | --- |
|  |  | Average | 19.7 | 16.1 | 81.5 |
|  |  | Standard deviation | 10.0 | 8.2 | - |
| Gatineau | PCS |  | TS (g/L) | VS (g/L) | % VS |
|  |  | Average | 24.0 | 21.8 | 90.8 |
|  |  | Standard deviation | 11.9 | 9.6 | - |

**Table S3:**
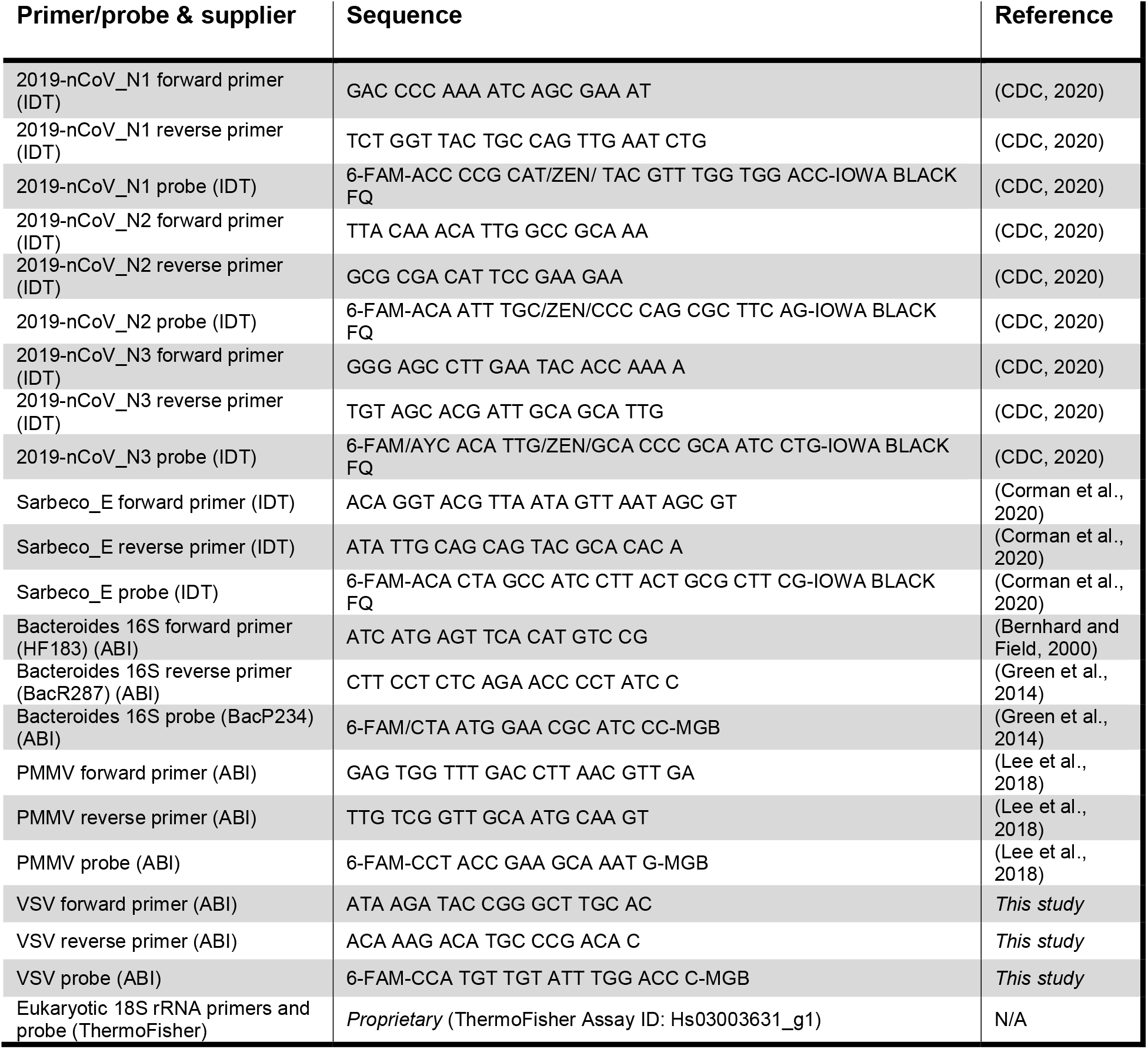
List of PCR primer and probe sets.

